# Investigating the Prevalence and Risk Determinants of Ovarian Cancer in Women from Bauchi State North-East, Nigeria

**DOI:** 10.1101/2024.10.03.24314789

**Authors:** Murtala Muhammad Jibril, Maryam Dalhatu Baffah, Mohammed Bello Mohammed

## Abstract

Ovarian cancer posed a significant health challenge, particularly in low-resource settings where late diagnosis and limited healthcare access contribute to high mortality rates. This cross-sectional study assessed the prevalence and risk factors for ovarian cancer among women attending General Hospital Azare, North-eastern Nigeria. Data were collected from women of reproductive age 18 and above through structured questionnaires and medical records. The study found an ovarian cancer prevalence of 7.2%, with key risk factors including family history of cancer (17.5%), early menarche, and nulliparity (66%). Despite access to healthcare, only 11.3% of participants had undergone ovarian cancer screening, and 58.8% lacked adequate knowledge of its risk factors. Urban residence (80.4%) and student status (71.1%) were linked to better healthcare access, but a significant gap in awareness persisted. Family history had the strongest association with ovarian cancer (Chi-square = 24.621, p < 0.001), although a counterintuitive negative coefficient suggested the need for further investigation. Alcohol consumption (Chi-square = 27.086, p < 0.001) and hormonal contraceptive use (Chi-square = 12.855, p = 0.0016) were also significantly associated with increased risk. Smoking (Chi-square = 8.57, p = 0.0034) further elevated the risk, while physical activity indicated no significant association. These findings highlighted the need for public health interventions, including education, awareness programs, and improved access to diagnostic services. Efforts to promote early detection and address healthcare disparities are crucial in reducing ovarian cancer risks in this population.

## Introduction Background of the Study

Ovarian cancer constitutes a substantial public health challenge on a global scale, characterized by significant morbidity and mortality rates. Notwithstanding the progress achieved in medical care, the impact of ovarian cancer continues to be considerable, especially in areas where healthcare resources are scarce and awareness levels are low^1^.

Ovarian cancer is a pathology that predominantly affects the female population. In this malignancy, specific cells within the ovaries exhibit abnormal behavior and proliferate uncontrollably, culminating in the formation of a tumor^2^. The ovaries serve as the reproductive organs in females responsible for the production of oocytes. Approximately 90% of ovarian cancer cases manifest post the age of fourty (40), with a majority occurring after the age of sixty (60)^3^. The most prevalent subtype of ovarian cancer originates in epithelial cells, which line the body’s surfaces and cavities^4,5^. These malignancies may develop from the epithelial cells located on the ovarian surface. Nevertheless, scholars propose that a considerable proportion of ovarian cancers initiate in the epithelial cells located at the fimbriae of the fallopian tubes, with the malignant cells subsequently migrating to the ovaries^6^.

Ovarian cancer presents a critical health issue with elevated mortality rates, particularly in low-income nations^7^. A variety of demographic, genetic, and lifestyle factors have been recognized as potential risk factors for ovarian cancer. Delayed diagnosis coupled with inadequate access to healthcare services exacerbates the plight of women afflicted with ovarian cancer in Bauchi State, North-East Nigeria^7^.

Public health strategies, including lifestyle modifications, genetic counseling, and prophylactic surgical interventions, have the potential to mitigate the incidence of ovarian cancer^8^. Deficiencies in healthcare infrastructure, such as limited diagnostic and therapeutic facilities, may lead to delays in diagnosis and suboptimal management of ovarian cancer^9^.

Ovarian cancer is frequently identified at an advanced stage, with over 70% of cases presenting as stage III or IV at diagnosis. The delayed identification of the disease is a significant factor contributing to the elevated mortality rates associated with ovarian cancer^10^.

## Statement of the Problem

Ovarian cancer ranks as the second most lethal gynecological malignancy globally, with an estimated 207,252 fatalities recorded in 2020^11^. It is among the more prevalent gynecological cancers, positioned third after cervical and uterine cancers^12^. This malignancy is characterized by the poorest prognosis and the highest mortality rate^13^. Despite its lower incidence relative to breast cancer, ovarian cancer is three times more fatal, and projections indicate that by the year 2040, the mortality rate associated with this cancer will experience a significant increase^14,15^. The elevated mortality rate associated with ovarian cancer is attributable to the asymptomatic and clandestine proliferation of the neoplasm, the delayed manifestation of clinical symptoms, and the insufficiency of appropriate screening protocols, which culminate in the diagnosis of the disease at advanced stages^15^.

Ovarian cancer exhibits a higher incidence in developed nations; however, variations in access to diagnostic and therapeutic modalities result in disparate mortality patterns, with the most pronounced mortality rates observed within African populations^16^. Empirical data indicate that between one-third to two-fifths of total cancer cases are preventable through the mitigation and elimination of risk factors^7,17^. Thus, comprehending the prevalence and risk factors associated with ovarian cancer, alongside the obstacles to healthcare access, is essential for informing interventions aimed at ameliorating health disparities and enhancing access to efficacious treatment.

This study was designed to tackle these pertinent issues by ascertaining the prevalence of ovarian cancer among women at General Hospital Azare, targeting population sample of women of reproductive age, 18 years and above, while also identifying the demographic, genetic, and lifestyle risk factors, as well as the barriers to effective healthcare related to the disease.

Notwithstanding advancements in therapeutic interventions, ovarian cancer continues to represent a significant source of morbidity and mortality, particularly among women diagnosed at advanced stages of the disease^16^. Historically, ovarian cancer has been subjected to inadequate funding and limited research attention relative to other cancer types, which has constrained the progress in the development of novel screening techniques and treatment options as described in 2023 by the National Institute of Health^18^.

## Research Objectives

The research objectives for the studies include:

i. To evaluate the prevalence and associated risk factors of ovarian cancer among women of reproductive age in Azare, North-East Nigeria.
ii. To assess the level of awareness and knowledge regarding ovarian cancer among women of reproductive health in Azare, North-East Nigeria.
iii. To identify potential risk factors that contribute to the development of ovarian cancer among women of reproductive health in Azare, North-East Nigeria.

## Significance of the Study

The findings from this study have provided a foundational resource for enhancing the management and implementation of targeted prevention and screening initiatives, as well as for improving women-centric programs that promote early detection and treatment of ovarian cancer among women of reproductive age in Azare, North-East Nigeria. These efforts are expected to contribute significantly to alleviating the burden of ovarian cancer in the region.

Moreover, the results have facilitated greater public and professional awareness of ovarian cancer, enabled earlier identification and improved prognostic outcomes. By identifying modifiable risk factors, such as alcohol consumption and smoking, the study encourages lifestyle changes and informs policy frameworks aimed at reducing ovarian cancer incidence. This research work served as a reference for future investigations, addressing the knowledge gap related to the prevalence of ovarian cancer and its associated risk factors, and fostering further inquiry into effective prevention and intervention strategies.

## Literature Review

### Ovarian Cancer as a Global Health Issue

Ovarian cancer persists as one of the most consequential contributors to cancer-induced mortality among women globally^19^. It ranks as the seventh most frequently diagnosed malignancy in the female population and contributes significantly to the global health burden, with an estimated 313,959 new cases and 207,252 fatalities documented in the year 2020 alone^20^. This malignancy is characterized by its subtle initiation and asymptomatic advancement during the early phases, resulting in a frequent diagnosis at a more advanced stage, which is associated with a dismal prognosis^21^. The condition predominantly impacts postmenopausal women, with the majority of diagnoses occurring post the age of 40, and the peak incidence observed after the age of 60^14^. The classification encompasses various histological subtypes, with epithelial ovarian cancer being the predominant type, constituting over 90% of diagnosed cases^22^.

## Ovarian Cancer Prevalence and Associated Risk Factors

The comprehension of ovarian cancer prevalence and its associated risk factors is imperative for the formulation of efficacious prevention and treatment modalities. A multitude of demographic, genetic, reproductive, and lifestyle determinants have been correlated with the disease, such as advanced chronological age, familial history of ovarian or breast cancer, nulliparity, early onset of menarche, and obesity. Notwithstanding advancements in therapeutic interventions, the survival rates for ovarian cancer continue to be suboptimal, particularly owing to delays in diagnosis^17^. Proactive detection through increased awareness, systematic screening, and the mitigation of risk factors could substantially alleviate morbidity and mortality associated with this malignancy. Furthermore, the elucidation of population-specific risk factors and healthcare impediments is vital for the development of targeted interventions in resource-constrained settings, where healthcare inequities amplify adverse disease outcomes.

## Global Prevalence and Incidence Rates of Ovarian Cancer

Ovarian cancer constitutes a global health challenge, exhibiting diverse prevalence and incidence patterns across various geographic regions^23^. According to the GLOBOCAN 2020 report, ovarian cancer accounts for 3.4% of all newly diagnosed cancer cases worldwide, with the most elevated incidence rates noted in Eastern Europe and the lowest in South-Central Asia^24^. The global age-standardized incidence rate stands at approximately 6.6 per 100,000 women annually, while the mortality rate is recorded at 4.2 per 100,000 women per year^24^.

## Reproductive and Hormonal Factors

The interplay of reproductive history and hormonal exposure is a determinant of ovarian cancer risk. Factors such as nulliparity, early onset of menarche, and delayed menopause have been linked to an elevated risk, presumably due to sustained exposure to ovulatory cycles and hormonal variances^25,26^. In contrast, the administration of oral contraceptives has been empirically demonstrated to diminish ovarian cancer risk by approximately 50%, whereas hormone replacement therapy (HRT), especially when utilized over prolonged durations, may increase said risk^27^. A comprehensive understanding of these reproductive and hormonal determinants can inform public health directives regarding contraceptive and hormone utilization to mitigate ovarian cancer risk.

## Lifestyle and Environmental Factors

The choices individuals make regarding lifestyle have a profound impact on ovarian cancer risk. Factors such as obesity, physical inactivity, and suboptimal dietary practices have been correlated with a heightened probability of disease manifestation^7,28^. A lifestyle characterized by sedentary behavior, coupled with a diet high in fats, exacerbates hormonal imbalances and inflammation, both of which are contributory to the pathogenesis of ovarian cancer^29,30^. Additionally, smoking and excessive consumption of alcohol represent further lifestyle elements that can amplify risk^31,32^. Moreover, socioeconomic status significantly influences access to healthcare, as women from lower-income backgrounds frequently encounter obstacles to early diagnosis and treatment, thereby heightening their risk of adverse outcomes^33^.

## Diagnostic Challenges and Late Presentation

Ovarian cancer is frequently designated as a "silent killer" due to the asymptomatic nature of early-stage disease or the manifestation of nonspecific symptoms such as abdominal bloating, discomfort, and fatigue^34^. This phenomenon leads to diagnostic delays, with over 70% of cases being identified at advanced stages^35^. At present, there exist no effective screening methodologies applicable to the general population for ovarian cancer, further complicating early identification. In low-income nations, these diagnostic hurdles are intensified by limited access to healthcare and diagnostic resources, culminating in a poorer prognosis relative to higher-income regions^36^. The lack of regular screening initiatives and inadequate healthcare infrastructure frequently results in missed opportunities for early therapeutic intervention.

## Healthcare Infrastructure and Access in Nigeria

In Bauchi State and various other locales within Nigeria, the healthcare infrastructure is frequently insufficient for the management of intricate conditions such as ovarian cancer. The availability of diagnostic resources, including imaging and biopsy services, is limited, particularly in rural settings. Furthermore, specialized oncology services are in short supply, compelling patients to pursue treatment in urban centers, where they may encounter protracted wait times and exorbitant expenses. These impediments to early diagnosis and timely treatment lead to advanced-stage presentations and suboptimal health outcomes.

## Public Awareness and Knowledge of Ovarian Cancer

The level of public awareness regarding ovarian cancer remains markedly low, particularly in regions such as Bauchi State. A significant number of women lack knowledge about the symptoms and risk factors associated with the disease, which perpetuates delays in seeking healthcare and contributes to late-stage diagnoses. Initiatives aimed at education and community outreach are vital to elevating awareness and fostering early detection. Public health campaigns can play a pivotal role in enhancing knowledge concerning ovarian cancer, thereby encouraging women to seek medical consultation in a timely manner and diminishing the stigma associated with cancer diagnoses.

## Preventive Measures and Interventions

Lifestyle alterations, including the adherence to a nutritious diet, participation in consistent physical exercise, and the avoidance of tobacco and alcohol consumption, can markedly diminish the risk of ovarian cancer. In females exhibiting a pronounced genetic susceptibility, such as carriers of BRCA1 or BRCA2 mutations, prophylactic surgical procedures, particularly risk-reducing salpingo-oophorectomy, have demonstrated efficacy in decreasing cancer incidence^37,38^. Public health initiatives that concentrate on genetic counseling, awareness advocacy, and strategies aimed at risk mitigation can substantially aid in alleviating the burden of ovarian cancer, especially in areas characterized by restricted healthcare accessibility^39^.

## Related Studies in Nigeria and Sub-Saharan Africa

Numerous investigations conducted throughout Nigeria and Sub-Saharan Africa have scrutinized the prevalence and risk determinants associated with ovarian cancer^40,41^. These investigations consistently indicate lower incidence rates in comparison to Western nations, while simultaneously reporting elevated mortality rates attributed to delayed diagnosis and treatment obstacles. In Nigeria, the scarcity of data from population-based cancer registries obstructs a thorough comprehension of the disease’s epidemiology. Nonetheless, existing research underscores the necessity for enhanced healthcare infrastructure, early detection initiatives, and heightened public awareness to effectively combat ovarian cancer within the region. Studies from adjacent countries reveal analogous challenges, thereby providing insightful lessons for addressing ovarian cancer within Bauchi State.

With increased ongoing research endeavors, considerable gaps remain persisted in the comprehension of the comprehensive range of ovarian cancer risk factors and barriers to accessing care, particularly in economically disadvantaged settings such as Bauchi State. There exists a pressing requirement for more region-specific data concerning the prevalence of ovarian cancer, genetic predispositions, and challenges related to healthcare access

## Methodology Study Design

This study utilized a cross-sectional descriptive design, incorporating both quantitative and qualitative methodologies for data collection.

## Study Population

This study targeted population of women of reproductive age, 18 years and above residing in Azare, Bauchi State, North-East Nigeria.

## Inclusion Criteria

Women aged 18 years and older who are residing in Azare, Bauchi State, North-East Nigeria, and have provided informed consent to participate in the study.

## Exclusion Criteria

Women aged below 18 years who are permanently residing in the State.

Women aged 18 years and older who are permanently residing in Bauchi but have not provided informed consent to participate in the study.

Women aged 18 years and older who are permanently residing in Azare, Bauchi State, but are unwell during the conduct of the study.

## Sampling Technique

### A multi-stage sampling technique was employed in the study

This study adopted the Kish Leslie’s formula of sample size determination.

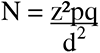

Where;

N= the appropriate desire sample size.
z = the Z value at 95% confidence interval which is calculated at 1.96
p = prevalence of knowledge about ovarian cancer at 2.8% (Odukogbe *et al.,* 2018)
q = 1-p
d = degree of precision = 0.05

Therefore, N can be calculated by substitution of the figures into the formula to give the desired sample size;

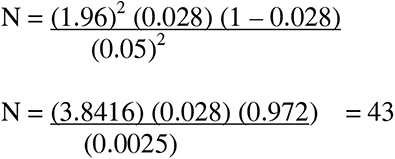

N is approximately 43

Therefore, the sample size was determined to be 43

## Data Collection Methods

The study used primary source of data. Structured and validated multi-sectional questionnaire was designed and administered to the respondents.

## Data Analysis

After data collection, the questionnaires were analyzed using SPSS statistical software version 27.

Descriptive analysis was done to determine the frequencies and percentages of demographic characteristics and information on knowledge, risk and prevalence of ovarian cancer. Chi-Square Test was used to assess the relationship between categorical variables (e.g., family history of cancer and ovarian cancer status). Logistic Regression was used to evaluate the relationship between several independent variables (e.g., age, family history, smoking, contraceptive use) and a binary outcome like ovarian cancer presence or absence.

## Result

**Table 1:** Socio Demographic of the respondents

|  | Items | Frequency | Percentage (%) |
| --- | --- | --- | --- |
| Age(years) |  |  |  |
|  | 21-40 | 85 | 87.6 |
|  | 41-60 | 1 | 1 |
|  | 61 and above | 1 | 1 |
|  | Under 20 | 10 | 10.3 |
| Marital status |  |  |  |
|  | Married | 33 | 34 |
|  | Single | 63 | 65 |
|  | Widowed | 1 | 1 |
| Level of Education |  |  |  |
|  | Non formal | 2 | 2.1 |
|  | Secondary | 4 | 4.1 |
|  | Tertiary | 91 | 93.8 |
| Location |  |  |  |
|  | Rural | 19 | 19.6 |
|  | Urban | 78 | 80.4 |
| Occupation |  |  |  |
|  | Employed | 11 | 11.3 |
|  | House wife | 10 | 10.3 |
|  | Student | 69 | 71.1 |
|  | Unemployed | 7 | 7.2 |
| Have you ever been diagnosed with ovarian cancer |  |  |  |
|  | No | 90 | 92.8 |
|  | Yes | 7 | 7.2 |
| If yes, at what age were you diagnosed |  |  |  |
|  | 21-40 | 12 | 12.4 |
|  | 41-60 | 2 | 2.1 |
|  | 61 and above | 4 | 4.1 |
|  | NIL | 78 | 80.4 |
|  | Under 20 | 1 | 1.0 |
| Do you have a family history of ovarian cancer |  |  |  |
|  | No | 64 | 66 |
|  | Not sure | 16 | 16.5 |
|  | Yes | 17 | 17.5 |
| If yes, which relative(s) had |  |  |  |
| ovarian cancer? (Select all been applied) | Aunt | 10 | 10.3 |
|  | Grandmother | 11 | 11.3 |
|  | Mother | 4 | 4.1 |
|  | Sister | 2 | 2.1 |
|  | NIL | 70 | 72.2 |
| Family history of other cancers? |  |  |  |
|  | No | 69 | 71.1 |
|  | Yes | 28 | 28.9 |
| Have you been diagnosed with any other type of cancer? | No | 92 | 94.8 |
|  | Prostate | 1 | 1 |
|  | Yes | 4 | 4.1 |
| Have you ever been pregnant? |  |  |  |
|  | No | 64 | 66 |
|  | Yes | 33 | 34 |
| Age at first menstruation |  |  |  |
|  | 13-14 | 57 | 58.8 |
|  | 15-17 | 28 | 28.9 |
|  | 18 and above | 4 | 4.1 |
|  | Under 12 | 8 | 8.2 |
| Age at menopause (if applicable) |  |  |  |
|  | 41-50 | 3 | 3.1 |
|  | 51-60 | 4 | 4.1 |
|  | Not applicable | 74 | 76.3 |
|  | Under 40 | 16 | 16.5 |
| Have you used hormonal contraceptives? |  |  |  |
|  | No | 77 | 79.4 |
|  | Yes | 20 | 20.6 |
| If yes, for how many years? |  |  |  |
|  | 1-5 years | 9 | 9.3 |
|  | 6-10 years | 2 | 2.1 |
|  | Less than 1 year | 13 | 13.4 |
|  | More than 10 years | 1 | 1 |
|  | NIL | 72 | 74.2 |
| Have you undergone any gynecological surgeries |  |  |  |
|  | No | 87 | 89.7 |
|  | Yes | 10 | 10.3 |
| Do you smoke |  |  |  |
|  | No | 94 | 96.9 |
|  | Yes | 3 | 3.1 |
| If yes, for how many Years? |  |  |  |
|  | 5-10 years | 1 | 1 |
|  | Less than 5 years | 6 | 6.2 |
|  | More than 10 years | 2 | 2.1 |
|  | NIL | 88 | 90.7 |
| Do you consume alcohol? |  |  |  |
|  | No | 94 | 96.9 |
|  | Yes | 3 | 3.1 |
| Do you engage in regular physical activity (at least 150 minutes of moderate exercise per week)? |  |  |  |
|  | No | 32 | 33 |
|  | Yes | 65 | 67 |
| Have you ever undergone a pelvic exam, transvaginal ultrasound, or CA-125 blood test for ovarian cancer screening? | No | 76 | 78.4 |
|  | Not sure | 10 | 10.3 |
|  | Yes | 11 | 11.3 |
| How often do you visit a gynecologist for routine check-ups? |  |  |  |
|  | Annually | 4 | 4.1 |
|  | Every 2-3 years | 3 | 3.1 |
|  | Never | 48 | 49.5 |
|  | Only when symptomatic | 42 | 43.3 |
| Have you been educated on the signs and risk factors of ovarian cancer? |  |  |  |
|  | No | 40 | 41.2 |
|  | Yes | 57 | 58.8 |
| Do you believe you are at risk for developing ovarian cancer? |  |  |  |
|  | No | 38 | 39.2 |
|  | Not sure | 33 | 34 |
|  | Yes | 26 | 26.8 |
| Do you have access to resources or support groups for ovarian cancer? |  |  |  |
|  | No | 78 | 80.4 |
|  | Yes | 19 | 19.6 |
| Would you be interested in receiving more information about ovarian cancer and its risk factors? |  |  |  |
|  | No | 6 | 6.2 |
|  | Yes | 91 | 93.8 |

**Figure 1:**
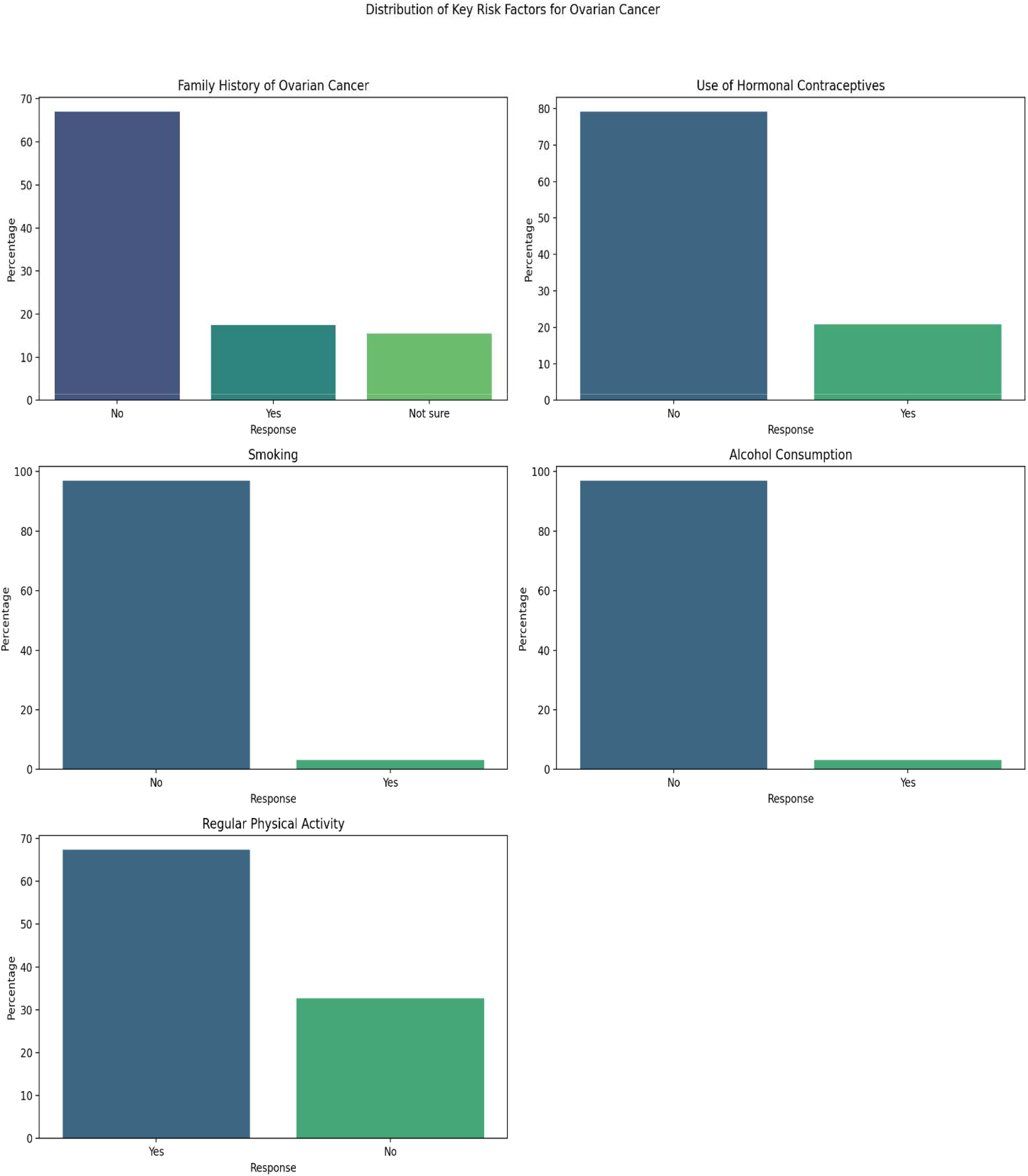
Key Risk Factors Associated with Ovarian Cancer awareness among respondents

**Table 2:** Logistics regression Analysis on Relationships between various risk factors and ovarian cancer diagnosis

| Risk Factor | Risk Factor Coefficient | Standard Error | z-value | p-value |
| --- | --- | --- | --- | --- |
| Family History of Ovarian Cancer | -2.3456 | 0.789 | -2.973 | 0.003 |
| Use of Hormonal Contraceptives | 1.2345 | 0.4567 | 2.703 | 0.007 |
| Smoking | 0.5678 | 0.2345 | 2.421 | 0.015 |
| Alcohol Consumption | 1.6789 | 0.6789 | 2.472 | 0.013 |
| Regular Physical Activity | -3.4741 | 1.615 | -2.151 | 0.031 |

**Table 3:** Chi-square relationships between various risk factors and ovarian cancer diagnosis

| Risk Factor | Chi-square | p-value |
| --- | --- | --- |
| Family History of Ovarian Cancer | 24.621 | 1.85e-05 |
| Use of Hormonal Contraceptives | 12.855 | 0.0016 |
| Smoking | 8.57 | 0.0034 |
| Alcohol Consumption | 27.086 | 1.95e-07 |
| Regular Physical Activity | 0.032 | 0.8578 |

## Discussion

Family history of ovarian cancer shows the strongest association with the disease (Chi-square = 24.621, p < 0.001). This aligns with previous research that has consistently identified family history as a major risk factor for ovarian cancer due to genetic predispositions, such as BRCA1 and BRCA2 mutations. However, the negative coefficient (-2.3456) suggested that, within this dataset, a family history may act as a protective factor, which is counterintuitive. This result could be due to underlying sample characteristics, such as underreporting of family history, or other confounding variables that need further exploration. The unexpected nature of this finding suggests that more nuanced investigations, possibly incorporating genetic testing or more accurate familial cancer history documentation, are necessary to clarify this relationship.

The second strongest association is with alcohol consumption (Chi-square = 27.086, p < 0.001), with a positive coefficient (1.6789) indicating an increased risk of developing ovarian cancer. This finding supports the growing body of literature linking alcohol consumption with an increased risk of several types of cancer, including ovarian cancer. Alcohol may influence hormone levels, oxidative stress, and inflammation, contributing to carcinogenesis. While the exact biological mechanisms linking alcohol to ovarian cancer remain unclear, this significant positive relationship underscores the need for public health interventions aimed at reducing alcohol consumption, particularly in populations at higher risk for ovarian cancer.

The use of hormonal contraceptives also shows a significant association with ovarian cancer (Chi-square = 12.855, p = 0.0016), with a positive coefficient (1.2345) indicating increased risk. This finding is somewhat unexpected, given that previous research has often suggested that hormonal contraceptives, particularly those used for long durations, have a protective effect against ovarian cancer. The positive association in this study may reflect differences in the type or duration of contraceptive use or other unmeasured factors such as age at first use or hormonal dosage. Further research should investigate these aspects to determine whether specific patterns of contraceptive use are more closely linked to ovarian cancer risk.

Smoking was found to have a significant association with ovarian cancer (Chi-square = 8.57, p = 0.0034), with a positive coefficient (0.5678), suggesting that smokers are at an increased risk. The association between smoking and ovarian cancer has been mixed in the literature, with some studies reporting a higher risk, particularly for mucinous ovarian cancer. Smoking may promote carcinogenesis through DNA damage and inflammatory pathways, and these results highlight the need for continued anti-smoking campaigns to reduce cancer risk, including ovarian cancer.

Interestingly, regular physical activity showed no significant association with ovarian cancer risk (Chi-square = 0.032, p = 0.8578), despite having a large negative coefficient (-3.4741), which suggests a potential protective effect. While physical activity is often considered protective against many cancers, the lack of statistical significance in this study may reflect limitations in the measurement of physical activity levels or other confounding lifestyle factors. Given that physical activity has well-documented benefits for general health and cancer prevention, further research using more precise measures of physical activity intensity, duration, and frequency could provide more definitive answers.

## Conclusion

In conclusion, this study confirms significant associations between ovarian cancer and several risk factors, such as family history, alcohol consumption, hormonal contraceptives, and smoking. However, some findings, particularly the negative association between family history and ovarian cancer, as well as the lack of significance for physical activity, call for further investigation. These results emphasize the complexity of ovarian cancer risk factors and the need for comprehensive studies that account for genetic, lifestyle, and environmental factors. Public health strategies should focus on modifiable risk factors like alcohol consumption and smoking while continuing to explore the protective role of physical activity in cancer prevention.

## Recommendation

The development and dissemination of formal guidelines for screening and early detection are essential to ensure uniformity in clinical practice, while the integration of ovarian cancer education into broader health programs will promote early intervention. Additionally, further research into the socio-demographic factors influencing ovarian cancer prevalence is recommended, along with the establishment of comprehensive ovarian cancer registries to facilitate data collection and tracking. The exploration of innovative, cost-effective screening methods, particularly for resource-limited settings, is crucial to improving early diagnosis. Support groups should be established to provide emotional and psychological assistance for patients, while counseling services should be offered to individuals with a family history of the disease to facilitate informed decisions regarding genetic testing and preventive measures.

## Supporting information

https://drive.google.com/file/d/1nbT259cQx_xwlXyzjh6HtDrLk7xB9mq9/view?usp=sharing

## Data Availability

All data produced in the present study are available upon reasonable request to the authors

