## Supplementary material for "Investigating the Prevalence and Risk Determinants of Ovarian Cancer in Women from Bauchi State North-East, Nigeria": https://drive.google.com/file/d/1nbT259cQx_xwlXyzjh6HtDrLk7xB9mq9/view?usp=sharing

Murtala Muhammad Jibril

Department of Human Anatomy, Faculty of Basic Medical Sciences, Bauchi State University, Bauchi State Nigeria

Maryam Dalhatu Baffah

Department of Human Anatomy, Faculty of Basic Medical Sciences, Bauchi State University, Bauchi State Nigeria

Mohammed Bello Mohammed

Department of Human Anatomy, Faculty of Basic Medical Sciences, University of Jos, Plateau State Nigeria

Corresponding Author:

Murtala Muhammad Jibril

Running Title:

Prevalence and Risk Determinants of Ovarian Cancer in Bauchi State

Word Count: 5216
