## Supplementary material for "Investigating the Prevalence and Risk Determinants of Ovarian Cancer in Women from Bauchi State North-East, Nigeria": https://drive.google.com/file/d/1nbT259cQx_xwlXyzjh6HtDrLk7xB9mq9/view?usp=sharing

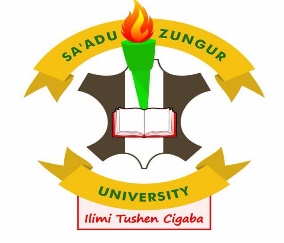

FACULTY OF BASIC MEDICAL SCIENCES, SA’ADU

ZUNGUR UNIVERSITY BAUCHI STATE

ETHICAL CLEARANCE CERTIFICATE

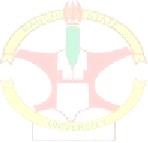

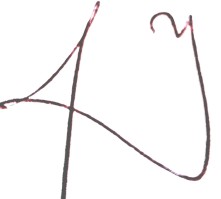

**GADAU**

Ref. No. BASUG/FBMS/REC/ VOL. 7/0018

**Date:** 26^th^ January, 2024

**Project Title:** INVESTIGATING THE PREVALENCE AND RISK DETERMINANTS OF OVARIAN CANCER IN WOMEN FROM BAUCHI STATE NORTH-EAST, NIGERIA

**Nature of the Project:** Research

**Principal Researcher:** Murtala Muhammad Jibril

**Co-**researcher: Maryam Dalhatu Bappah, Mohammed Bello Mohammed

Faculty of Basic Medical Sciences’ Research and Ethics Committee (FBMSREC) gives this approval in respect of the undertakings contained in the above-mentioned project and research instrument(s). Should any other instrument(s) be used, this requires separate authorization. Therefore, the researcher may commence with the research as from the date of this certificate, with reference to the number indicated above.

You will be expected to provide progress report upon completion of your study.

**SIGNED**

**MUSLIM ABBA Ph.D**

**CHAIRMAN REC**

**26th January, 2024**
